# Leveraging neuroscience education to address stigma related to opioid use disorder (OUD) in the community: A pilot study

**DOI:** 10.1101/2023.12.11.23299824

**Authors:** Evan J. Kyzar, Melissa Arbuckle, Adam Abba-Aji, Krishna Balachandra, Joseph Cooper, Adriane Dela Cruz, Ellen Edens, Brady Heward, Michael Jibson, Ayana Jordan, Daniel Moreno-De-Luca, Hannah Pazderka, Mohit Singh, Jeremy Weleff, Bernice Yau, Justin Young, David A. Ross, National Neuroscience Curriculum Initiative (NNCI)

## Abstract

Opioid use disorder (OUD) and overdose deaths are a public health crisis. One contributing factor is stigma towards people who use opioids. We developed and conducted a public-facing, half-day educational event designed to challenge misperceptions about OUD from a contemporary neuroscience perspective. Participants engaged with three different resources on the neurobiology of addiction; at the end of the event, they rated its effectiveness. We also collected and compared pre- and post-event composite OUD stigma scales. Participants rated our approach and the overall event as highly effective. Additionally, OUD stigma scores were lower immediately following the event, and this decrease was primarily driven by decreased internalized stigma. Here, we demonstrate an effective proof-of-concept that an accessible, public-facing, neuroscience education event may reduce OUD stigma in the community.

## Introduction

North America is in the midst of an opioid crisis, with fatal opioid overdoses reaching over 75,000 per year in 2020 (Humphreys et al., 2022) and rising. This crisis has had devastating effects across socioeconomic groups, with disproportionate impacts on historically marginalized communities due to inequitable systems of care (Friedman & Hansen, 2022). There is an urgent need for interventions that address opioid use disorder (OUD) not only with patients and their family members, but also in the broader community.

Multiple factors have contributed to the current crisis, including the proliferation of synthetic opioids, limited access to evidence-based interventions for OUD, and social determinants of health. Another key contributor is the insidious role of stigma, in large part the legacy of moral conceptualizations that invite blame and prejudice towards persons struggling with addiction. For individuals with OUD, internalized negative beliefs may limit their engagement with healthcare systems (Chang et al., 2019; Olsen & Sharfstein, 2014), leading to poorer treatment outcomes. Similarly, stigma among clinicians, especially those without specialty training in substance use disorders (SUDs), may prevent them from implementing evidence-based approaches if they subconsciously blame patients or otherwise misunderstand SUDs, treatment, and recovery (Klusaritz et al., 2023; Stone et al., 2021). In the general population, increased stigma towards people who use opioids is associated with lower levels of support for addiction services (Pyra et al., 2022). Despite this, relatively few studies have directly tested the impact of internalized or perceived stigma on treatment outcomes in OUD patients (Crapanzano et al., 2019).

One approach to addressing stigma is to combat ignorance and fear with understanding and hope – as has happened in other branches of medicine, such as cancer (Else-Quest & Jackson, 2014). For many years, our understanding of the biology of SUDs was limited by available scientific approaches. Only recently has modern neuroscience enabled a robust understanding of addiction: how both genetic and environmental factors can translate into vulnerability or resilience through shared neurobiological pathways. As our understanding of addiction shifts, an opportunity exists for educational interventions that harness these new findings in a way that can decrease stigma and enhance engagement with treatment. While there have been a small number of studies on various methods to decrease stigma related to addiction (Livingston et al., 2012), to our knowledge there have been no studies evaluating the effect of neuroscience education on OUD stigma.

To this end, we designed a public-facing educational intervention that was rooted in principles of adult learning. We piloted the approach in a community sample that included individuals with lived experience, family members, and health care providers. Our primary objective was to determine the feasibility and effectiveness of our event. We compared pre- and post-event stigma scores retrospectively to assess the impact on participant attitudes.

## Methods

### Event recruitment

Participants were recruited via flyers in the local area, outreach to community organizations, and social media. We advertised that we were developing an educational program on OUD and wanted feedback from people with lived and living experience, family members, and the broader public. Following standards for community engagement, participants were compensated for their time (with a 100 CAD gift card) and breakfast and lunch were provided. Participants were informed that their feedback would be used for quality improvement.

### Educational event design and data collection

The 3-hour educational event was held on March 20^th^, 2023 at the University of Alberta in Edmonton, Canada. The program was designed around principles of adult learning theory: using experiential learning approaches, facilitating differentiation (consistent with a constructivist model), leveraging social connection among participants, and incorporating formative assessment tools (Bransford et al., 2000; Handelsman et al., 2004). While we focused on neuroscience resources, our event offered a holistic message of hope and recovery – that existing treatments work and that recovery is possible. The neuroscience resources were designed according to best practices of effective scientific communication (e.g., using narrative approaches to make content broadly accessible, connecting content to real-world scenarios of lived experience, and attending to data visualization).

Participants filled out a pre-survey that included: whether they or their family member(s) had lived experience with addiction and whether they worked in health care (in order to maintain anonymity and encourage participation, we minimized the number of questions asked and did not collect any individually identifying data); five Likert scale questions derived from the Opening Minds Stigma Scale (Modgill et al., 2014) modified for a community sample; and open-ended responses to prompts following three case vignettes (Supplemental Methods).

Participants were assigned to five breakout rooms of ∼10 participants with 2-3 facilitators per room. For each of the three vignettes (Fig. 1A) participants worked in groups of 2-3 to discuss their initial responses to the scenario, review a neuroscience-focused educational resource, and then reflect on how the resource might change the way they thought about the scenario. The resources were crafted to highlight core questions – and misconceptions – relating to OUD: 1) a video on genetic and environmental contributions to risk (*Jam Jar - Opioid Use Disorder*, 2022. https://vimeo.com/706154349); 2) a short article on long-term affective changes that contribute to relapse after abstinence (Bommersbach et al., 2020); and 3) a video on the contribution of negative affective states to opioid use (Kyzar, 2022. https://nncionline.org/course/dr-evan-kyzar-the-hidden-side-of-addiction/). Following each small group activity there was a brief full group discussion with summary of key themes.

**Figure 1.**
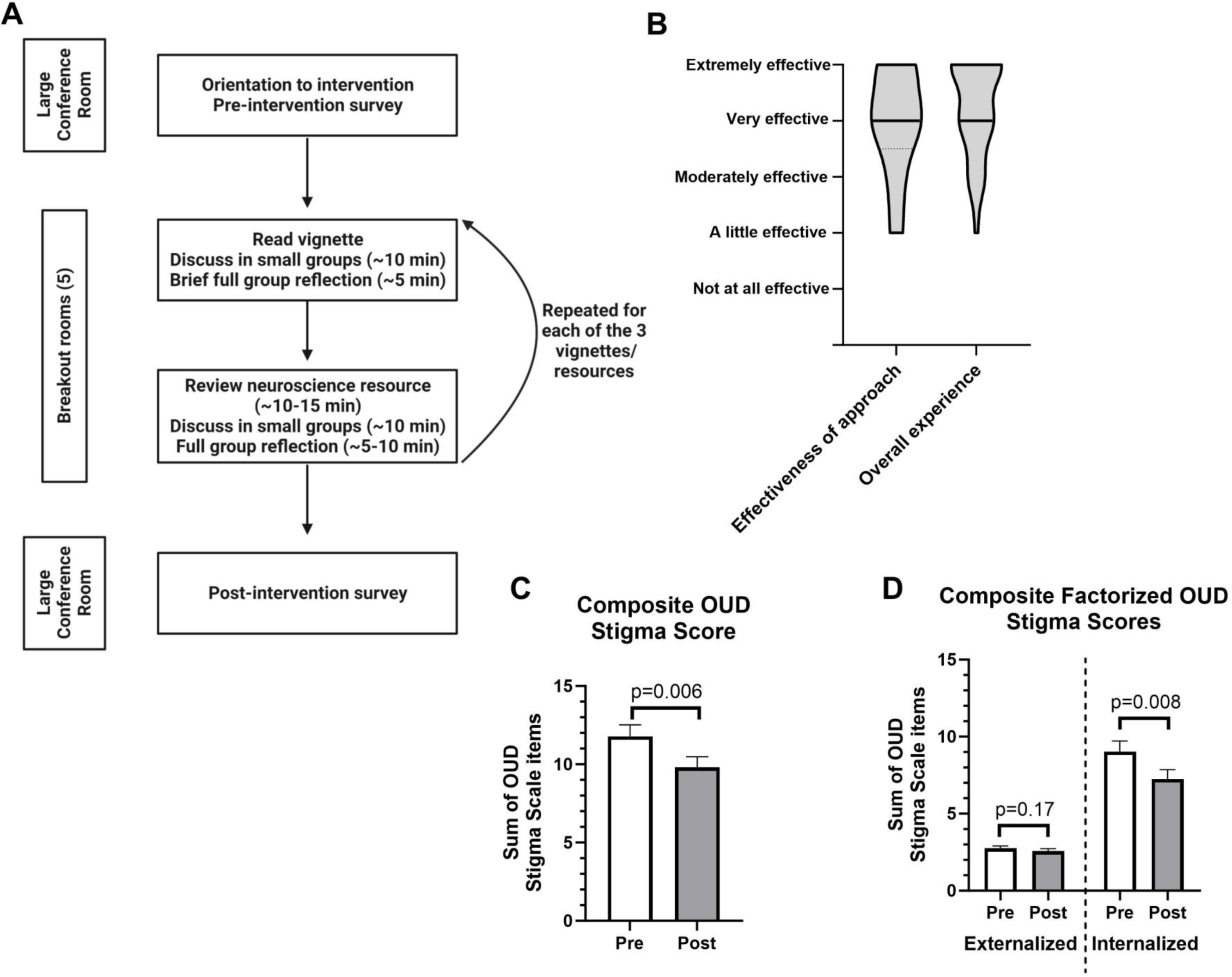
Design of a neuroscience-focused educational event and its perceived effectiveness and impact on stigma scales related to opioid use disorder (OUD). A) Flowchart outlining the organization of the program. B) Data on the perceived effectiveness of the approach and the overall experience of the event, rated on a Likert scale which was numerically converted for graphical representation as follows: Extremely effective = 5; Very effective = 4; Moderately effective = 3; A little effective = 2; Not at all effective = 1. Graphs show smoothed violin plots with black bars indicating median response. C) Pre- and post-event ratings for composite OUD stigma score: Z=-2.74, W=-240, *p*= 0.006 (primary hypothesis tested at α=0.05). Graph shows mean ± SEM. D) Pre- and post-event ratings for two summed OUD stigma scale factors (termed externalized and internalized) identified using exploratory factor analysis (see Fig. S1); Externalized factor (2 questions): Z=-1.44, W=-44, *p*=0.017; Internalized factor (3 questions): Z=-2.65, W=-185, *p*=0.008 (significant when Bonferroni corrected at α=0.025). Graph shows mean ± SEM. All stigma scales were rated on a Likert scale which was numerically converted for statistical analyses as follows: Strongly agree = 5; Agree = 4; Neither agree nor disagree = 3; Disagree = 2; Strongly disagree = 1. All data were analyzed using Wilcoxon signed-ranks tests with n=37, as the Likert data was not normally distributed. See Supplemental Table 1 for raw means of pre- and post-event data. Note that the post-event survey was completed approximately 3-3.5 hours after the pre-event survey, immediately following the conclusion of the educational event.

Immediately after the breakout room activities, participants were asked to complete a survey that included the same questions as the pre-survey plus additional questions about the effectiveness of our approach and their overall experience of the event (Supplemental Methods).

### Statistical analyses and primary hypothesis

Our data analyses were approved by the Research Ethics Board (REB) at the University of Alberta. Likert scale responses were converted to numerical data. We examined the correlation between participants’ rating of the effectiveness of approach and overall experience using a Pearson test. We tested internal consistency of the five OUD stigma questions with Cronbach’s alpha and performed exploratory factor analysis. These data were summed to create a composite OUD stigma score. Our main hypothesis related to retrospective analysis of OUD stigma scores was that the program would decrease composite stigma scores (α=0.05). We performed secondary analyses on the identified factors from the factor analysis (created by summing questions from each of the factors) and each stigma question with Bonferroni correction for multiple comparisons (α=0.025 for factors; α=0.01 for each question). We used nonparametric Wilcoxon signed-rank tests for these analyses. We also examined the change in composite OUD stigma score depending on answers to the demographic questions (by Mann-Whitney *U*-test) and depending on ratings of the effectiveness of approach and overall experience of the event (by one-way ANOVA). Data were analyzed in R v4.0.4 and GraphPad Prism 9.

## Results

### Participant ratings of effectiveness and overall experience

Overall, 47 unique participants from the community attended the event (Fig. 1A), and 37 participants completed both the pre- and post-event formative assessment (78.72%). Amongst participants who completed both surveys, 16/37 (43.24%) had lived experience with addiction, 24/37 (64.86%) had a family member with lived experience of addiction, and 17/37 (45.95%) were employed as a health care worker. These categories were not mutually exclusive, with 19/37 (51.35%) participants answering affirmatively to ≥2 of these categories and 3/37 (8.11%) answering in the negative to all. Participants found both the approach and the overall experience to be effective (Fig. 1B), and ratings of the effectiveness of our approach and the overall experience were highly correlated (R^2^ = 0.6492, *p*<0.0001).

### Effect on OUD stigma scores

We performed a retrospective analysis of stigma scores collected pre- and post-event. Internal consistency of our adapted stigma scale was adequate (Cronbach’s α = 0.75), and factor analyses revealed two factors related to externalized stigma (i.e., towards others) and internalized stigma (i.e., self-stigma), respectively (Fig. S1). For example, the prompt “I struggle to feel compassion for a person with opioid use disorder” was in the externalized factor, while “I would see myself as weak if I had opioid use disorder and could not fix it myself” was in the internalized factor. This factor structure was consistent with the identified factors of “Attitude” (similar to externalization of stigma) and “Disclosure/Help Seeking” (similar to internalization of stigma) from the Opening Minds Stigma Scale (Modgill et al., 2014).

We found that summed post-event composite OUD stigma scores were significantly lower compared to pre-event scores (Z=-2.74, W=-240, *p*=0.006) (Fig. 1C). We next performed secondary analyses to determine if particular factors or questions may be driving the decrease in post-event composite scores. Summed internalized stigma questions identified in the factor analysis were decreased post-event (Z=-2.65, W=-185, *p*=0.008) with no effect on questions related to externalized stigma (Fig. 1D). Raw mean stigma scores decreased for all five questions (Fig. S2; Supplemental Table 1). Interestingly, the two largest decreases in individual questions came on questions related to internalized or self-stigma (Fig. S2D-E), though neither met stringent criteria for statistical significance after correction for multiple comparisons. Raw Likert scale responses for pre- and post-event surveys are shown in Fig. S3.

### Effect of demographics on change in composite OUD stigma scores

We tested whether belonging to particular demographic groups had an effect on the change in composite OUD stigma scores between pre- and post-event surveys, finding that there were no differences between participants with and without lived experience (Fig. S4A), participants with and without family member(s) with lived experience (Fig. S4B), and health care workers versus non-health care workers (Fig. S4C). We found that the change in composite OUD stigma scores between pre- and post-event surveys did not depend on ratings of the overall experience (Fig. S4D) or the effectiveness of our approach (Fig. S4E).

## Discussion

Here we describe a proof-of-concept pilot of a public-facing, community-level program focused on addressing OUD stigma using neuroscience education. We designed our intervention with an explicit focus on adult learning theory (Bransford et al., 2000; Handelsman et al., 2004), utilizing accessible resources to create interactive learning opportunities. Given the pilot nature of our event, we were principally interested in individuals’ experience of the event. Participants viewed both our approach and the overall event as highly effective (Fig. 1B).

We performed a retrospective analysis of survey data to determine whether our event had an effect of OUD stigma. Many of the participants had personal experience with addiction, and stigma scores (particularly for externalized stigma) were relatively low in our sample. Despite this potential floor effect, mean composite OUD stigma scores decreased post-event (Fig. 1C), an effect which appeared independent of individuals’ prior experience with addiction and/or working in health care. Interestingly, the decrease in OUD stigma scores appears to have been driven by decreases in internalized or self-stigma (Fig. 1D).

Our pilot event and reported results add to the emerging literature on addressing stigma through educational approaches. While some authors have suggested that neuroscience education may paradoxically increase stigma (Buchman et al., 2011), our results suggest that, at least in our sample, this was not the case. There are a number of possible reasons why we observed a decrease in OUD stigma. First, while we highlighted accessible neuroscience resources, our event also conveyed a holistic message of hope that emphasized that effective treatments are available and that recovery is possible. Similar recovery-oriented interventions have been shown to reduce internalized stigma in mental illnesses (Kroska & Harkness, 2021). Additionally, our resources highlighted the combined influences of genetic factors and environmental stressors, as well as the importance of addressing modifiable risk factors in recovery. We speculate that the success of our approach hinged on this balance between calling attention to critical neuroscientific concepts while also conveying hope in a recovery-centered way.

We emphasize that our results represent a proof-of-concept pilot, and as such there are important caveats and limitations. Our sample size was limited, and the participatory nature of the event raises the possibility of sampling biases. This likely contributed to the lower pre-event stigma scores, though we saw a decrease in post-event scores regardless. We minimized questions on personal information to decrease barriers to participation, and it is possible that our intervention had differential effects on specific groups. Additionally, the exact psychological mechanisms that underlie the observed decrease in OUD stigma are unclear. Others factors that may contribute include the impact of increased social support and connection with other participants with similar backgrounds (Heilig et al., 2016; Judd et al., 2023), the above-mentioned hopeful and recovery-oriented message, and the use of personal stories from those with lived experience, which have been separately shown to decrease stigma (Kissell et al., 2022).

As this was a community-facing pilot study, future work should attempt to extend these findings to a broader and more heterogenous sample, including individuals with higher levels of baseline stigma. Additionally, though individuals with lived experience did attend our event, this was explicitly not a clinical population. Neuroscience education for other chronic illness such as chronic pain improves functional outcomes (Louw et al., 2011), and future studies should directly address whether a similar educational strategy may alter treatment outcomes in clinical OUD populations.

## Conclusions

There are numerous strategies that could help combat the ongoing opioid crisis, including improved access to evidence-based treatments, robust harm reduction approaches, and initiatives to address underlying social determinants of health. Novel educational interventions, such as the one outlined in this report, could offer a powerful additional tool by offering a message of hope and recovery, decreasing stigma, enhancing engagement with evidence-based treatments, and facilitating more thoughtful public policies.

## Data Availability

All data produced in the present study are available upon reasonable request to the authors

## Disclosures and acknowledgements

The NNCI is funded in part by the Deeda Blair Research Initiative Fund for Disorders of the Brain through support to the Foundation for the National Institutes of Health, with additional funding from the Society of Biological Psychiatry (SOBP) and the American College of Neuropsychopharmacology (ACNP). EJK is supported by a training grant through the National Institute of Mental Health (R25 MH086466) and the Leon Levy Fellowship in Neuroscience. MA is supported by the above-named National Institute of Mental Health grant (R25 MH086466). AA-A is supported by grants from the New Frontiers in Research Fund, PRIHS-6, and Mitacs Accelerate fund. DM-D-L is supported by CASA Mental Health as the CASA Research Chair. DAR is supported by the Alberta Health Services Chair in Mental Health Research. The authors report no biomedical financial interests or potential conflicts of interest related to the present work.

---

**Figure S1.**
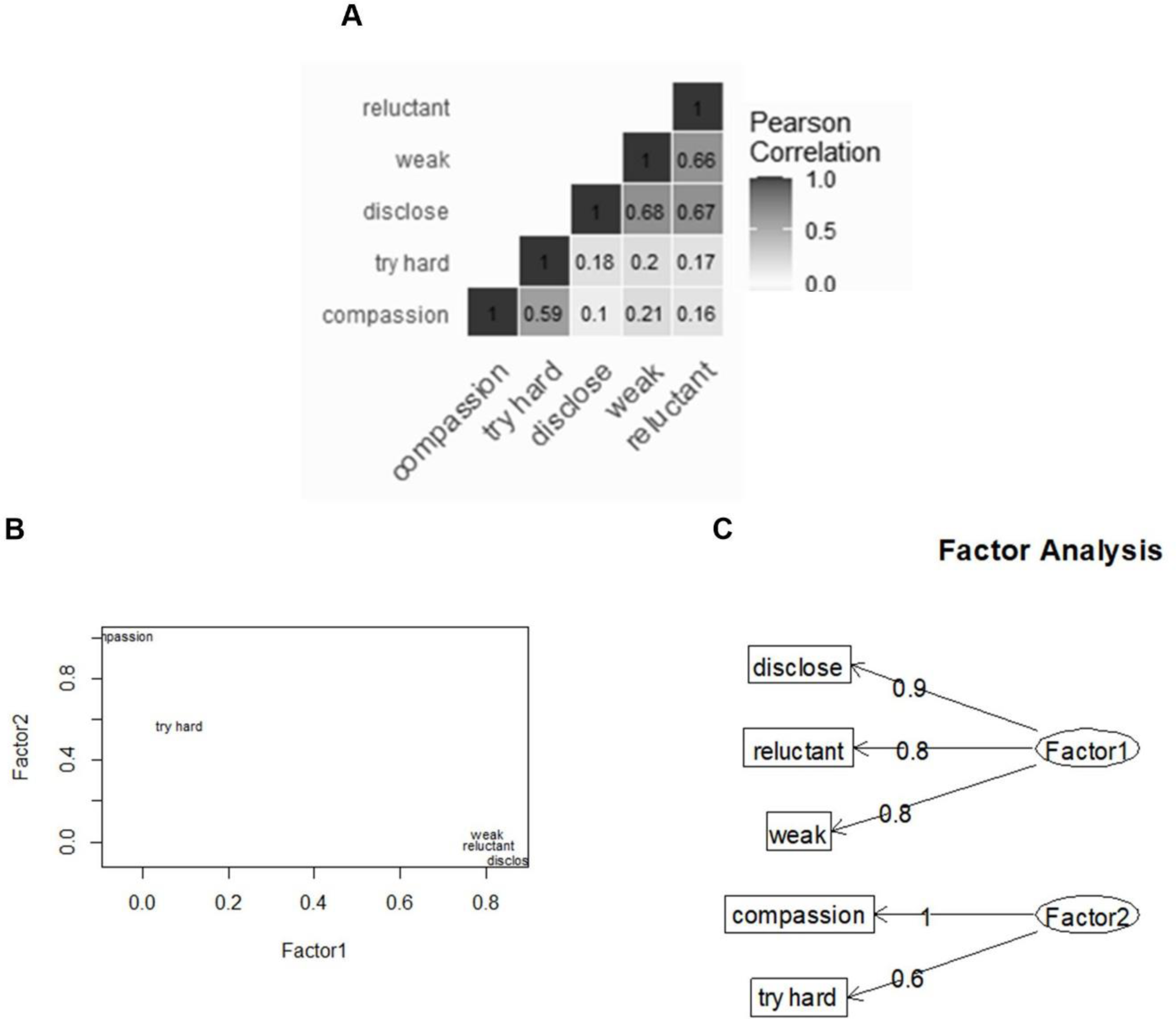

**Figure S2.**
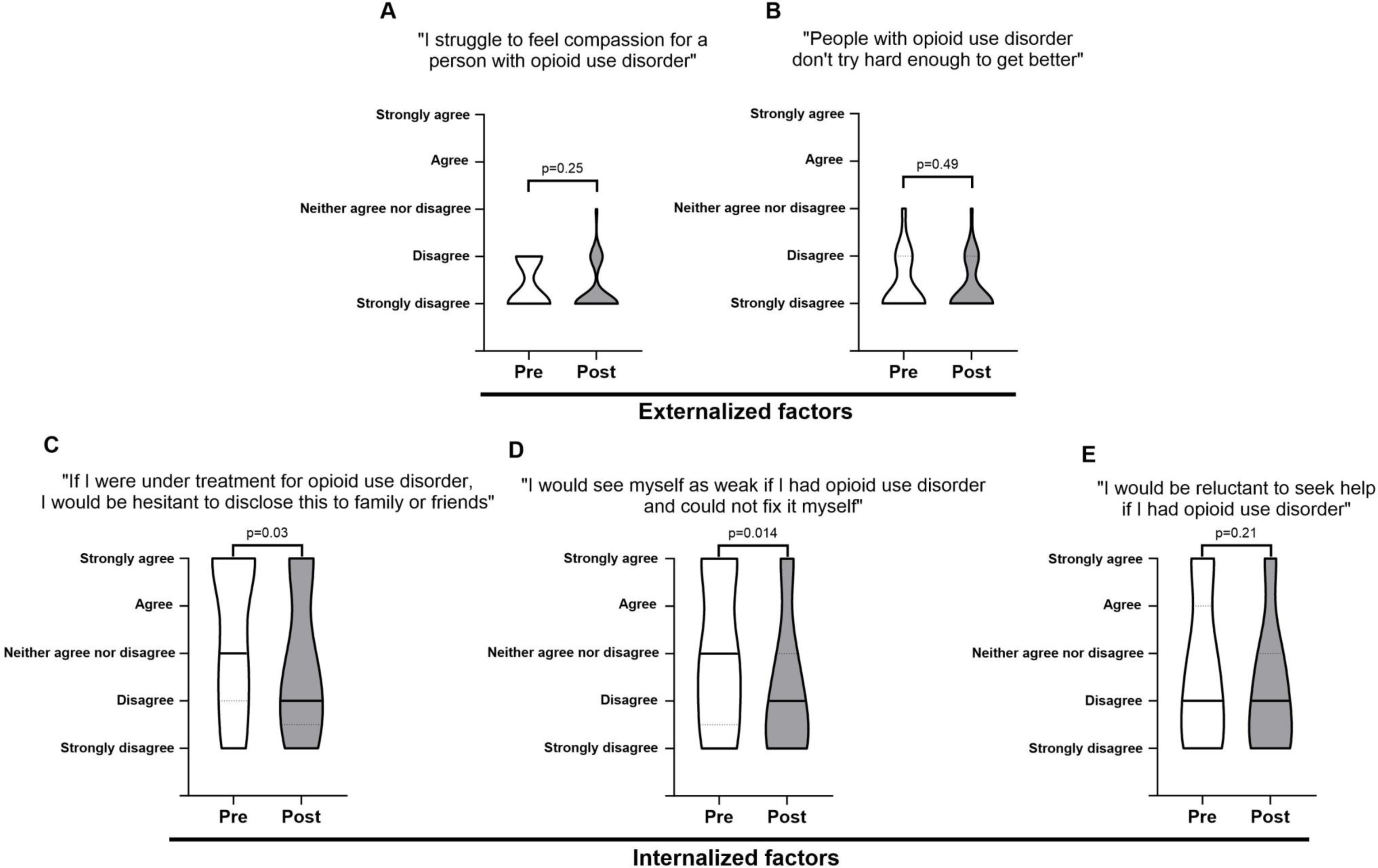

**Figure S3.**
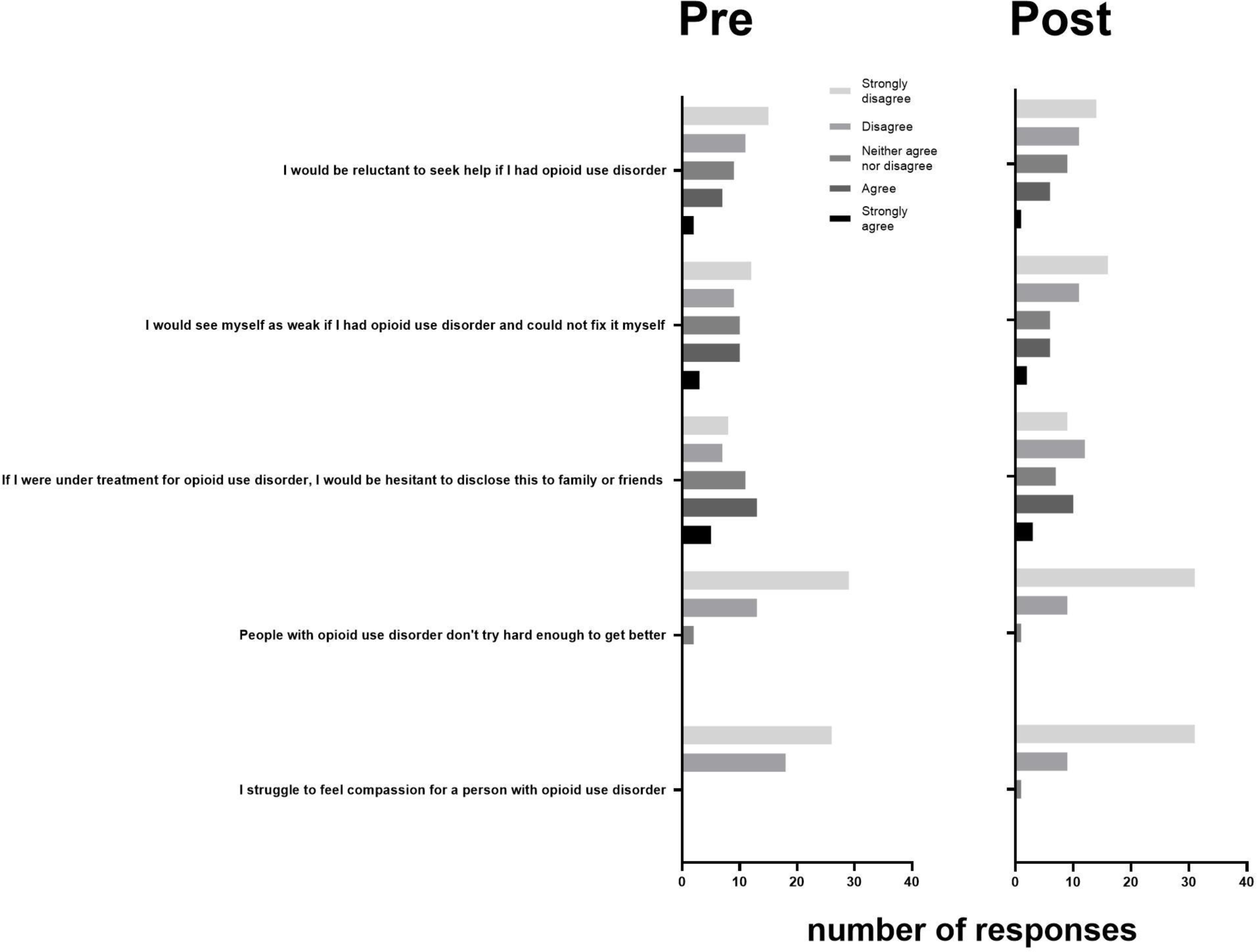

**Figure S4.**
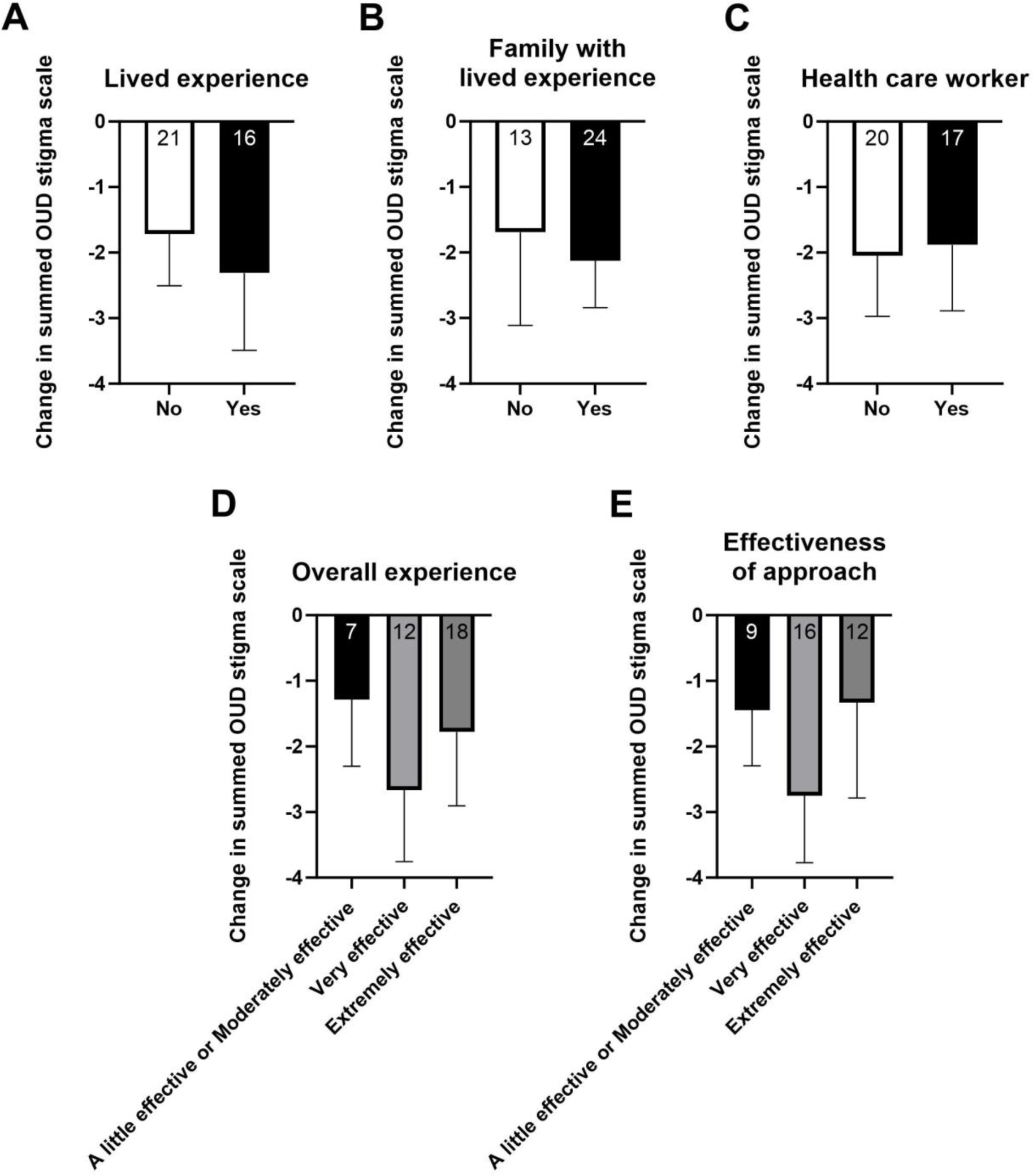

## Notes

### Competing Interest Statement

The authors have declared no competing interest.

### Author Declarations

Research Ethics Board of the University of Alberta gave ethical approval for this work

