## Supplemental Information for "Leveraging neuroscience education to address stigma related to opioid use disorder (OUD) in the community: A pilot study"

Kyzar et al.

***Supplemental Methods***

*Survey questions*

1. Please rate your level of agreement with the following statements: [I struggle to feel compassion for a person with opioid use disorder.][a†]

2. Please rate your level of agreement with the following statements: [People with opioid use disorder don't try hard enough to get better.][a†]

3. Please rate your level of agreement with the following statements: [If I were under treatment for opioid use disorder, I would be hesitant to disclose this to family or friends.][a†]

4. Please rate your level of agreement with the following statements: [I would see myself as weak if I had opioid use disorder and could not fix it myself.][a†]

5. Please rate your level of agreement with the following statements: [I would be reluctant to seek help if I had opioid use disorder.][a†]

6. Overall effectiveness of today's experience [How effective did you find this approach to education?][b‡]

7. Overall effectiveness of today's experience [How would you rate today's overall experience?][b‡]

[a]rated on 5-point Likert scale [numeric conversion in brackets] (Strongly agree [5]; agree [4]; neither agree nor disagree [3]; disagree [2]; strongly disagree [1])

[b]rated on 5-point Likert scale [numeric conversion in brackets] (Extremely effective [5]; Very effective [4]; Moderately effective [3]; A little effective [2]; Not at all effective [1])

[†]Asked both pre- and post-event

[‡]Asked only post-event

*Vignettes*

Vignette 1:

You come from the kind of small town where everyone knows everyone else's business. You start working at a shelter for individuals experiencing homelessness when you meet John. You've heard about John and his family from a distance for years. John has struggled with addiction since he was in high school and people have gossiped about him and judged him from afar. Some even blame John's parents – they say that his father used to use drugs and the two of them must have done something wrong to cause their son to become "an addict."

When you meet him, he talks about growing up in a small town, parents that worked too hard to make it by with too little, a father that was "too stern," and an older brother who's been successful and is now living in Toronto – the same kind of story that you've heard from other friends who grew up in that part of the province.

He starts crying when he talks about his addiction, then looks up and plaintively asks: "why me?"

Vignette 2:

You're visiting a close friend's family and learn that one of the boys didn't make it home for the holidays. Their mother tells you that "he just hasn't been the same since his car accident" and subsequent back surgery. They managed his post-operative pain with opioids but things never fully resolved. His family physician continued trying different opioid medications for more than a year but they never seemed to work well enough – in fact, his mom wonders if they made it worse. About six months ago, they stopped his opioids because they were worried about addiction, but things haven't gotten any better. He's always complaining about some kind of pain and the family can't shake the feeling that "something just isn't right – he seems angry all the time… he doesn't enjoy things the way he used to."

His mom seems tired and sad, frustrated by feeling helpless. She takes a deep breath and tries to compose herself. "I don't understand. The accident was two years ago – why can't he just get over it?"

Vignette 3:

You go to a coffee shop to meet your friend, Sarah. You hadn't heard from her in a long time until a couple of weeks ago when she reached out to connect. You know that she's been going through a lot – her mother died about a year ago and you imagine that she's been deeply affected by local news stories about residential schools.

When you sit down, she looks restless and uncomfortable. After a bit of general catching up, there's a lull in the conversation and Sarah becomes quiet. Her eyes dart around the room and she surprises you by disclosing that she's fallen into opioid addiction. She tells you about her experiences going through withdrawal - shaking, sweating, nausea, and the feeling that she is going to crawl out of her skin – and that she's terrified to go through that again. She describes a deep sense of shame and guilt and says that she's scared she is losing control. "I used to love the high but these days I don't even like how it makes me feel… so why can't I stop?"

*Supplemental Figures and Tables*

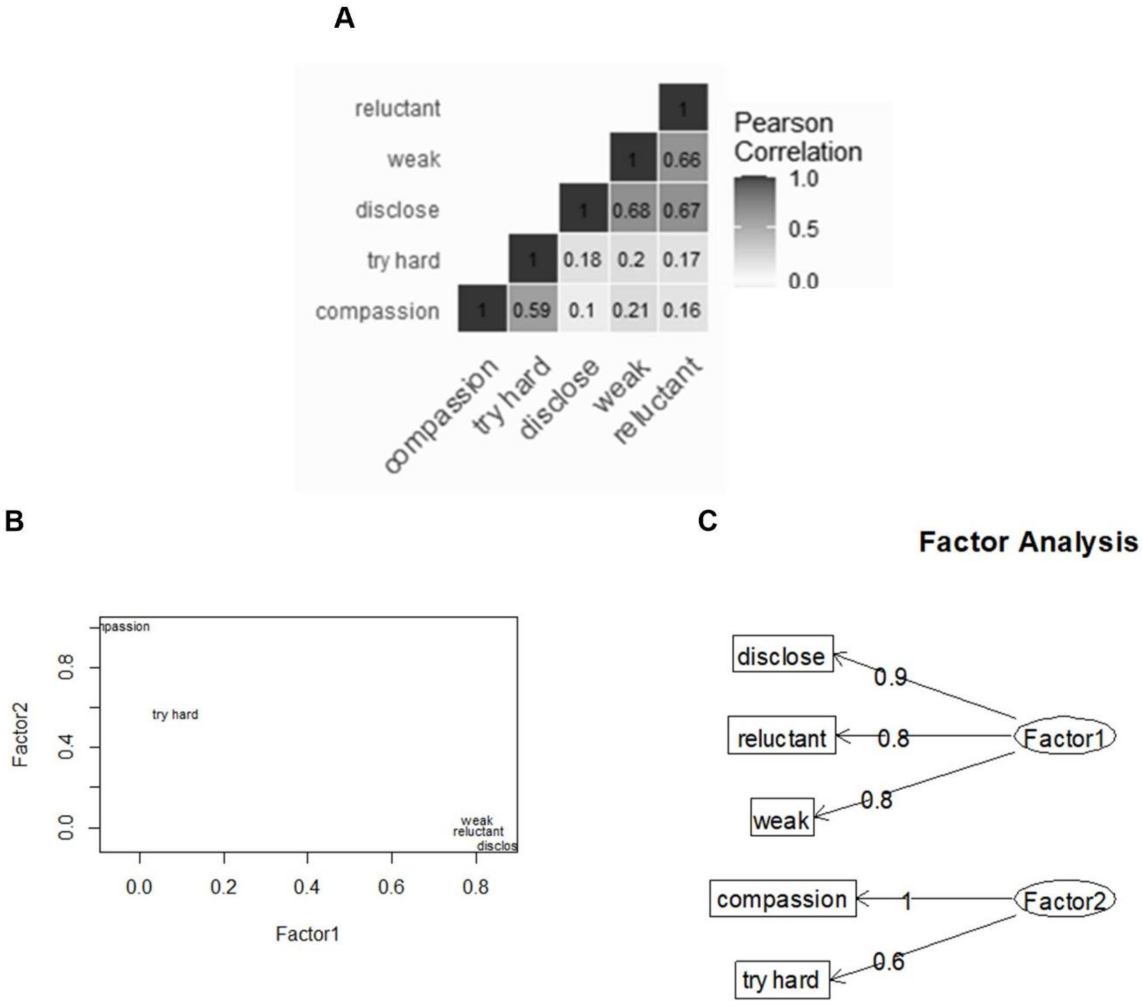


**Figure S1. Internal consistency, correlation, and factor analysis of opioid use disorder (OUD) stigma scale data used in this study.** A) Correlation matrix of Pearson's correlations between the 5 OUD stigma scale items, with within-square values representing $R^2$ values. B) Factor model plot with respective factor loadings on y-axis (externalized factors; Factor2) and x-axis (internalized factors; Factor1). C) Factor model flowchart showing factor loadings. In all graphs, *reluctant* = [I would be reluctant to seek help if I had opioid use disorder.]; *weak* = [I would see myself as weak if I had opioid disorder and could not fix it myself.]; *disclose* = [If I were under treatment for opioid use disorder, I would be hesitant to disclose this to family or friends.]; *try hard* = [People with opioid use disorder don't try hard enough to get better.]; *compassion* = [I struggle to feel compassion for a person with opioid use disorder.].

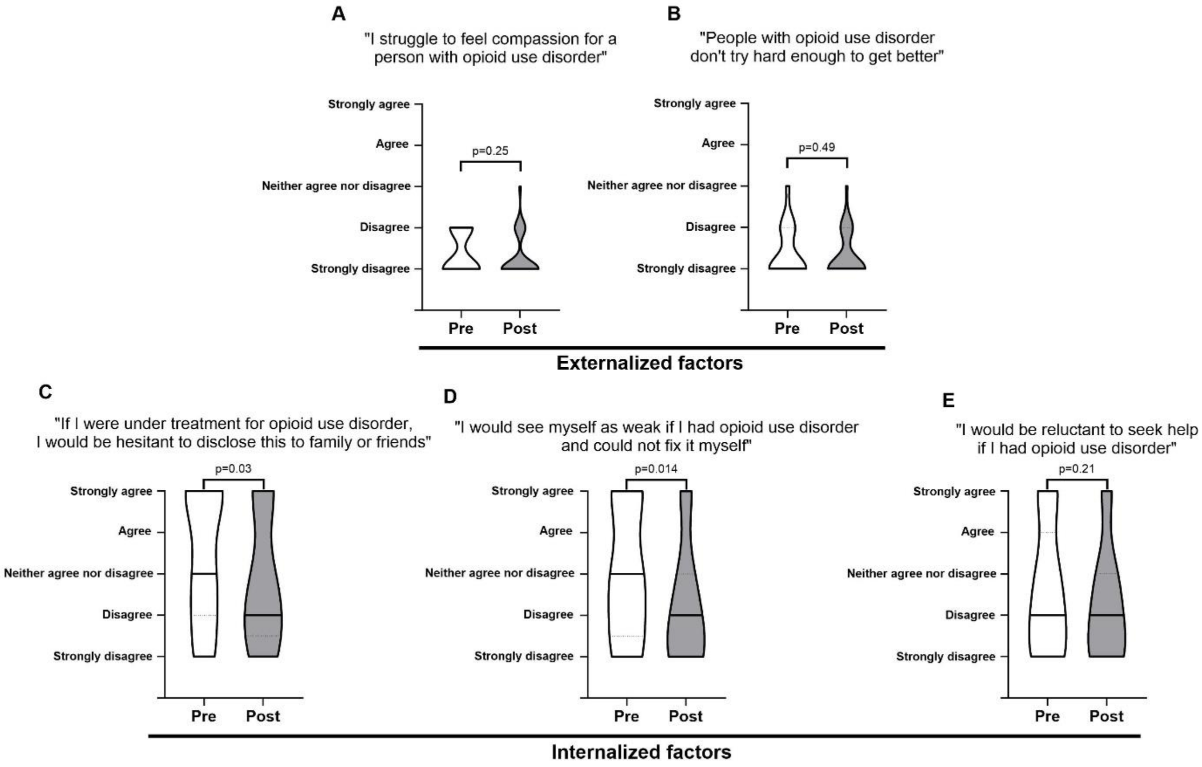


**Figure S2. Individual stigma score items related to opioid use disorder (OUD) compared pre- and post-event.**

A) Pre- and post-event ratings for query “I struggle to feel compassion for a person with opioid use disorder”: Z=-1.41, W=-18, *p*=0.025. Graph shows smoothed violin plots with black bars indicating median response. This question was included in the externalized factor.

B) Pre- and post-event ratings for query “People with opioid use disorder don’t try hard enough to get better”: Z=-0.83, W=-15, *p*=0.49. Graph shows smoothed violin plots with black bars indicating median response. This question was included in the externalized factor.

C) Pre- and post-event ratings for query “If I were under treatment for opioid use disorder, I would be hesitant to disclose this to family or friends”: Z=-2.20, W=-69, $p$= 0.03 (not significant when Bonferroni corrected at revised α=0.01). Graph shows smoothed violin plots with black bars indicating median response. This question was included in the internalized factor.

D) Pre- and post-event ratings for query “I would see myself as weak if I had opioid use disorder and could not fix it myself”: Z=-2.43, W=-119, $p$=0.014 (not significant when Bonferroni corrected at revised α=0.01). Graph shows smoothed violin plots with black bars indicating median response. This question was included in the internalized factor.

E) Pre- and post-event ratings for query “I would be reluctant to seek help if I had opioid use disorder”: Z=-1.30, W=-64, $p$=0.21. Graph shows smoothed violin plots with black bars indicating median response. This question was included in the internalized factor.

All stigma scales were rated on a Likert scale which was numerically converted for statistical analyses as follows: Strongly agree = 5; Agree = 4; Neither agree nor disagree = 3; Disagree = 2; Strongly disagree = 1. All data were analyzed using Wilcoxon signed-ranks tests with n=37, as the Likert data was not normally distributed. Lines and labels underneath the graphs represent each questions inclusion in the externalized or internalized factor following factor analysis. See Supplemental Table 1 for raw means of pre- and post-event data. Note that the post-event survey was completed approximately 3-3.5 hours after the pre-event survey, immediately following the conclusion of the educational event.

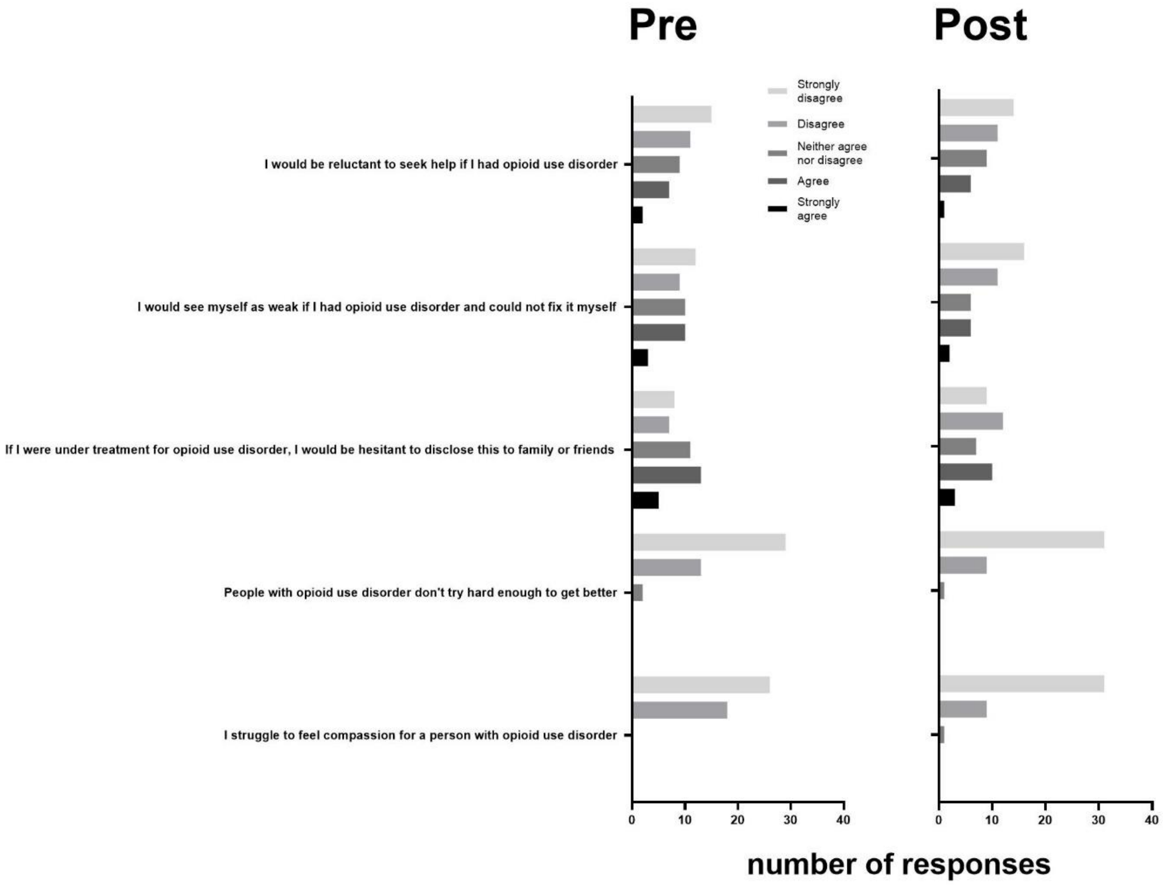


**Figure S3. Raw Likert scale responses to questions used for Figures 1 and S2**.

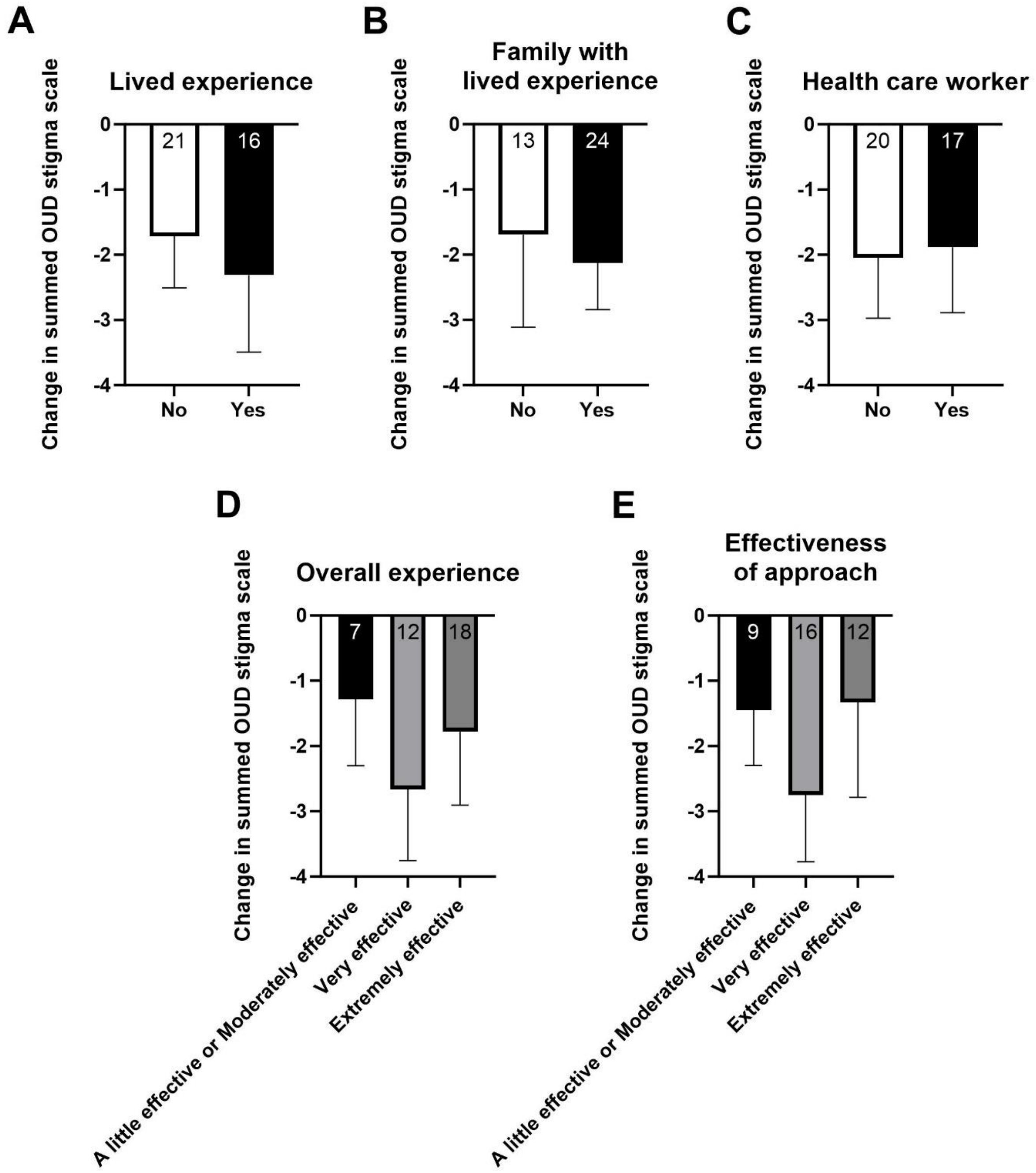


**Figure S4. Change in composite opioid use disorder stigma scale (OUD) by demographic information and event ratings**. There were no statistical differences in the change in composite OUD stigma score (calculated by post-event minus pre-event)

in A) participants with and without lived experience with addiction (*U*=160, T=303, *p*=0.99 by Mann-Whitney *U*-test); B) participants with and without family members with lived experience with addiction (*U*=137, T=267, *p*=0.54 by Mann-Whitney *U*-test); and C) health care workers versus non-health care workers (*U*=164, T=329, *p*=0.87 by Mann-Whitney *U*-test).

D) Change in composite opioid use disorder (OUD) stigma scale (calculated by post-event minus pre-event) grouped by perception of overall experience ($F_{(2, 34)}$ = 0.2818, *p*= 0.7562 by one-way ANOVA).

E) Change in composite opioid use disorder (OUD) stigma scale (calculated by post-event minus pre-event) grouped by perception of the effectiveness of approach ($F_{(2, 34)}$ = 0.4991, *p*= 0.6114 by one-way ANOVA).

Sample sizes are indicated on the respective graphs. Data is shown as mean ± standard error of the mean (SEM).

| Figure | Pre-event | Post-event |
|---|---|---|
| *Fig. 1C* | 11.784 ± 1.498 | 9.811 ± 1.352 |
| *Fig. 1D (Externalized)* | 2.757 ± 0.308 | 2.568 ± 0.320 |
| *Fig. 1D (Internalized)* | 9.027 ± 1.400 | 7.243 ± 1.253 |
| *Fig. S2A* | 1.378 ± 0.164 | 1.270 ± 0.169 |
| *Fig. S2B* | 1.378 ± 0.198 | 1.297 ± 0.173 |
| *Fig. S2C* | 3.405 ± 0.541 | 2.757 ± 0.512 |
| *Fig. S2D* | 3.000 ± 0.527 | 2.243 ± 0.474 |
| *Fig. S2E* | 2.622 ± 0.517 | 2.243 ± 0.477 |

**Supplemental Table 1. Raw mean values (± 95% confidence interval) summed, factorized, and individual stigma scale items**. All stigma scales were rated on a Likert scale which was numerically converted for statistical analyses as follows: Strongly agree = 5; Agree = 4; Neither agree nor disagree = 3; Disagree = 2; Strongly disagree = 1.
